# The physiological demands of Singing for Lung Health compared to treadmill walking

**DOI:** 10.1101/2020.12.08.20245746

**Authors:** Keir EJ Philip, Adam Lewis, Sara C Buttery, Colm McCabe, Bishman Manivannan, Daisy Fancourt, Christopher Orton, Michael I Polkey, Nicholas S Hopkinson

**Affiliations:** National Heart and Lung Institute, Imperial College London, London, United Kingdom; Department of Health Sciences, Brunel University, London, United Kingdom; Department of Behavioural Science and Health, University College London, London, United Kingdom; Royal Brompton and Harefield NHS Foundation Trust, London, United Kingdom

**Keywords:** singing, exercise, physical activity, physiology, heart rate, oxygen, ventilation, COVID-19

## Abstract

Participating in singing is considered to have a range of social and psychological benefits. However, the physiological demands of singing, whether it can be considered exercise, and its intensity as a physical activity are not well understood. We therefore compared cardiorespiratory parameters while completing components of Singing for Lung Health (SLH) sessions, with treadmill walking at differing speeds (2, 4, and 6km/hr). Eight healthy adults were included, none of whom reported regular participation in formal singing activities. Singing induced physiological responses that were consistent with moderate intensity activity (METS: median 4.12, IQR 2.72 - 4.78), with oxygen consumption, heart rate, and volume per breath above those seen walking at 4km/hr. Minute ventilation was higher during singing (median 22.42L/min, IQR 16.83 - 30.54) than at rest (11L/min, 9 - 13), lower than 6km/hr walking (30.35L/min, 26.94 - 41.11), but not statistically different from 2km/hr (18.77L/min, 16.89 - 21.35) or 4km/hr (23.27L/min, 20.09 - 26.37) walking. Our findings suggest the metabolic demands of singing may contribute to the health and wellbeing benefits attributed to participation. However, if physical training benefits result remains uncertain. Further research including different singing styles, singers, and physical performance impacts when used as a training modality is encouraged.

## Background

Singing is a ubiquitous cultural practice throughout history and across the world(1), and participation in singing is believed to have a range of health and wellbeing benefits(2, 3). Research to date has predominantly focused on psychosocial, and psychobiological impacts(4-8). However, the cardiorespiratory demands of singing, and the potential for it to serve as a form of exercise and contribute to daily physical activity are less well examined.

An appreciation of the physiological demands of singing could improve understanding of how best to use singing in a therapeutic capacity. An example of a structured therapeutic singing intervention is Singing for Lung Health (SLH), which has been developed as a strategy to help people with respiratory disease(8-12), particularly those who continue to be limited by breathlessness despite optimal medical care(13-15). Though high-quality research on the impacts of SLH is limited(16), participants report a range of biopsychosocial impacts(8, 17), including physical improvements relating to balance(18) and physical aspects of quality of life(8). Furthermore, it is known that exercise training is one of the most effective management strategies for people with long term respiratory conditions(19), usually in the form of pulmonary rehabilitation, however many people are unable to access PR(20), hence alternative approaches could be complementary in expanding provision of exercise training opportunities and diversifying delivery modalities, if an evidence base were to be established.

Additionally, identifying existing, enjoyable, and well attended physical activities of sufficient intensity to be considered exercise is useful from a public health and health promotion perspective. Physical activity is important both to maintain health and to mitigate the impact of long term medical conditions(21). This is particularly relevant during the present COVID-19 pandemic, where physical distancing measures to reduce risk of COVID-19 transmission, combined with the concerns about the virus itself, are having unintended negative impacts including inactivity, social isolation, and anxiety(22, 23). As such, there is an urgent need to provide and support evidence-based strategies, that are deliverable in the current situation and beyond, which could, for example include online singing groups(18, 24).

To evaluate this further we undertook a study to compare cardiorespiratory parameters during singing, and various SLH exercises, with i) rest and ii) three different walking speeds.

## Methods

### Participants

We conducted a non-blinded observational study. A convenience sample of colleagues and staff at the National Heart and Lung Institute (NHLI) were approached face-to-face and invited to participate in the study. The initial intention was to recruit 12 participants, however the implementation of restrictions on aerosol generating procedures due to the COVID-19 pandemic meant we decided to stop at eight. None of the participants sung regularly. Inclusion criteria included: age 18 to 99 years; no significant medical conditions or active musculoskeletal disease impairing exercise; no contraindications to exercise or spirometry as per American Thoracic Society/European Respiratory Society criteria; and capacity to consent to exercise testing.

The study was prospectively registered at clinicaltrial.gov (NCT04121351). Ethical approval was granted by the Imperial College Research Ethics Committee (IREC) (19IC5429). All participants provided informed written consent.

### Physiological parameter assessment

Physiological parameters assessed were VO_2_ ml/kg/min, end tidal CO_2_ (kPa), heart rate (bpm), minute ventilation (L/min), respiratory rate (breaths/min), mean volume per breath (L/breath). Gas analysis and flow were collected using JLab software package, Breath-by-Breath, and the Jaeger Oxycon Pro and Vyaire Oxycon mobile devices (see photos wearing device in supplementary information). The device was calibrated between participants as per standard protocol. Heart rate was assessed using the Polar heart rate monitor (Polar, Finland). Measures of perceived effort and dyspnoea were recorded at baseline and following each component according to the Borg RPE and Dyspnoea scales. Each stage of the protocol was completed for two minutes with 20 seconds between each section to allow for a verbal reminder of the next stage of the protocol to the participant, equipment check, and change of participant position if necessary. The two-minute duration of protocol components was selected based on a compromise between recommendations regarding exercise testing guidelines(25), being representative of real-world SLH sessions, and pilot work comparing the second minute values with longer protocol durations, which suggested stability of values during the second minute of each component. As such, the mean value from the second minute of assessment was used. Data were recorded continuously as the protocol was completed by each participant.

Spirometry was conducted as per American Thoracic Society/European Respiratory Society Guidelines (ATS/ERS) (26). Physical activity intensity was considered as light, moderate and vigorous, according to Metabolic Equivalents (METs), derived from the VO_2_ ml/kg/min data, with light physical activity if below 3 METS, moderate if between 3-6 METS and vigorous if above 6 METS(27). METs for each component were calculated by dividing by 3.95 ml/kg/min, which was the median measurement for the group during the resting phase 1.

### Singing Protocol

Singing for Lung Health (SLH) is a structured group singing programme for people with chronic respiratory conditions(8) see https://www.blf.org.uk/support-for-you/singing-for-lung-health. The components of a SLH session are similar to those found in most community choirs and singing groups, but in addition, aim improving participants symptoms through song, breathing exercises and relaxation techniques. We selected SLH components as an established method of group-singing for which the session content has been clearly defined and evaluated indicating intervention fidelity(8, 11). Each component was demonstrated by AL to each participant who briefly practiced the content of each component to show understanding, before resting for 30 mins during study set up.

The full study protocol is provided in the online supplementary material. However, components in brief were as follows:

1. Baseline assessment at rest
2. Physical warm up (gentle, dance-based, to music)
3. Rhythm exercise, seated singing
4. Pitch exercise, seated singing
5. Vocal fricatives, focusing on consonant vocalisations
6. Song repertoire, standing singing
7. Rest component 2
8. Treadmill walking 2km/hr
9. Treadmill walking 4km/hr
10. Treadmill walking 6km/hr

Walking speeds were selected as being representative of a slow, medium, and fast walk. These speeds also cover the NHS definition of a ‘brisk’ walk of 3 miles per hour (4.8km/hr)(28), recommended as moderate intensity exercise which can increase aerobic fitness(29).

Rest component two was included to ensure that the protocol included sufficient time for full recovery between components, and to enable participants physiological parameters to return to baseline before the walking components.

### Statistical analysis

Analyses were carried out using Stata 14 (StataCorp, TX). The Friedman test was used to assess for differences in the impact of protocol components on physiological parameters. Post-hoc Wilcoxon-signed rank tests were used for pair wise comparisons between singing, rest, and walking. Statistical significance was set at p<0.05. Readers wanting to adjust for multiple comparisons within each physiological parameter could apply a Bonferroni alpha of 0.013 (p<0.05 divided by 4 tests per parameter). Further adjustment for multiple comparisons across the different physiological parameters was not calculated given our sample size was small and our study exploratory. Data are presented to two significant figures.

## Results

Freidman tests demonstrated that the protocol components induced differences in all physiological parameters: VO_2_ ml/kg/min (Q (9) = 65.78, P <0.001); METs (Q (9) = 65.78, P <0.001); end tidal CO_2_ (Q (9) = 45.19, P <0.001); heart rate (Q (9) = 58.44, P <0.001); minute ventilation(Q (9) = 57.30, P <0.001; respiratory rate (Q (9) = 48.60, P <0.001); volume per breath (Q (9) = 43.31, P <0.001); Borg breathlessness scale (Q (9) = 32.91, P <0.001); Borg perceived exertion scale (Q (9) = 40.50, P <0.001).

Data are shown in Figure 1. The main singing condition (protocol component 6) showed that singing induced statistically significant increases in oxygen consumption, heart rate, and volume per breath compared with rest conditions, walking at 2km/hr, or walking at 4km/hr (pairwise comparisons using Wilcoxon-sign rank test). Minute ventilation was higher during the singing component than at rest, and lower than walking at 6km/hr, but not statistically significantly different from walking at 2 or 4km/hr. End tidal CO_2_ was higher singing than at rest or walking at 2km/hr, but not statistically different from walking at 4 or 6km/hr. Borg breathlessness scale ratings suggest singing was associated with an increased sensation of breathlessness compared with rest and all walking speeds. Perceived exertion during singing was greater than during rest and walking at 2km/hr, but not different from walking at 4 or 6km/hr. Respiratory rate was lower during singing than rest or walking, however this is likely due to the phrasing of the songs, rather than being a representative of a physiologically driven respiratory rate.

**Figure 1:**
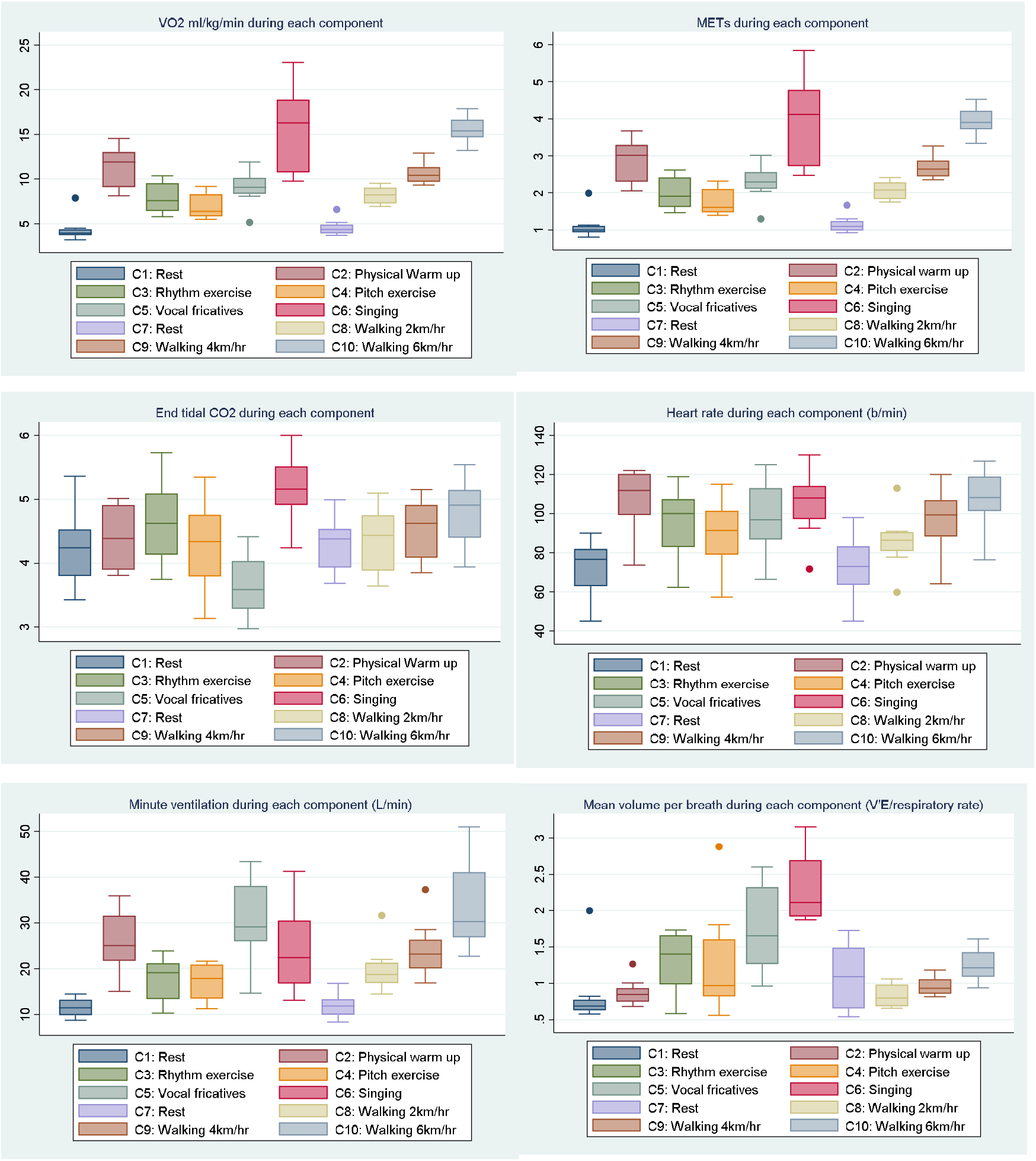
Box and whisker plots of physiological parameters during each component of the protocol. For box and whisker plots the line in the centre of the box represents the median, the box includes the first to third quartiles, the whiskers indicate upper and lower values (excluding outliers), the dots represent possible outliers.

**Figure 1: Box and whisker plots of VO**_**2**_ **ml/kg/min, heart rate, Minute ventilation, breathing frequency, and volume per breath (OR physiological variables assessed) during the protocol components. Freidman tests demonstrated that the protocol components induced differences in all physiological parameters P <0**.**001**

**Table 1:**
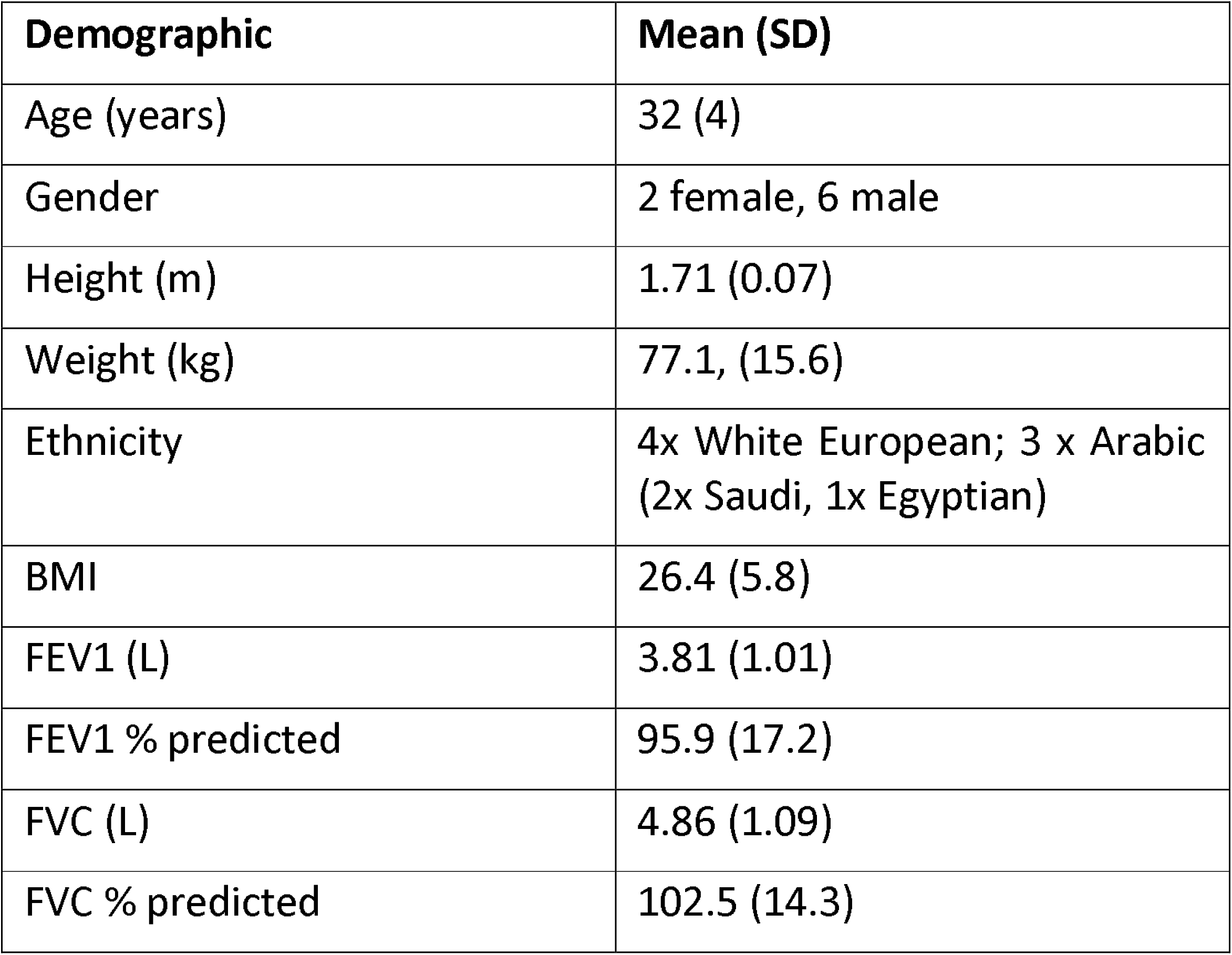
Participant Characteristics

| **Demographic** | **Mean (SD)** |
| --- | --- |
| Age (years) | 32 (4) |
| Gender | 2 female, 6 male |
| Height (m) | 1.71 (0.07) |
| Weight (kg) | 77.1, (15.6) |
| Ethnicity | 4x White European; 3 x Arabic<br>(2x Saudi, 1x Egyptian) |
| BMI | 26.4 (5.8) |
| FEV1 (L) | 3.81 (1.01) |
| FEV1 % predicted | 95.9 (17.2) |
| FVC (L) | 4.86 (1.09) |
| FVC % predicted | 102.5 (14.3) |

**Table 2:**
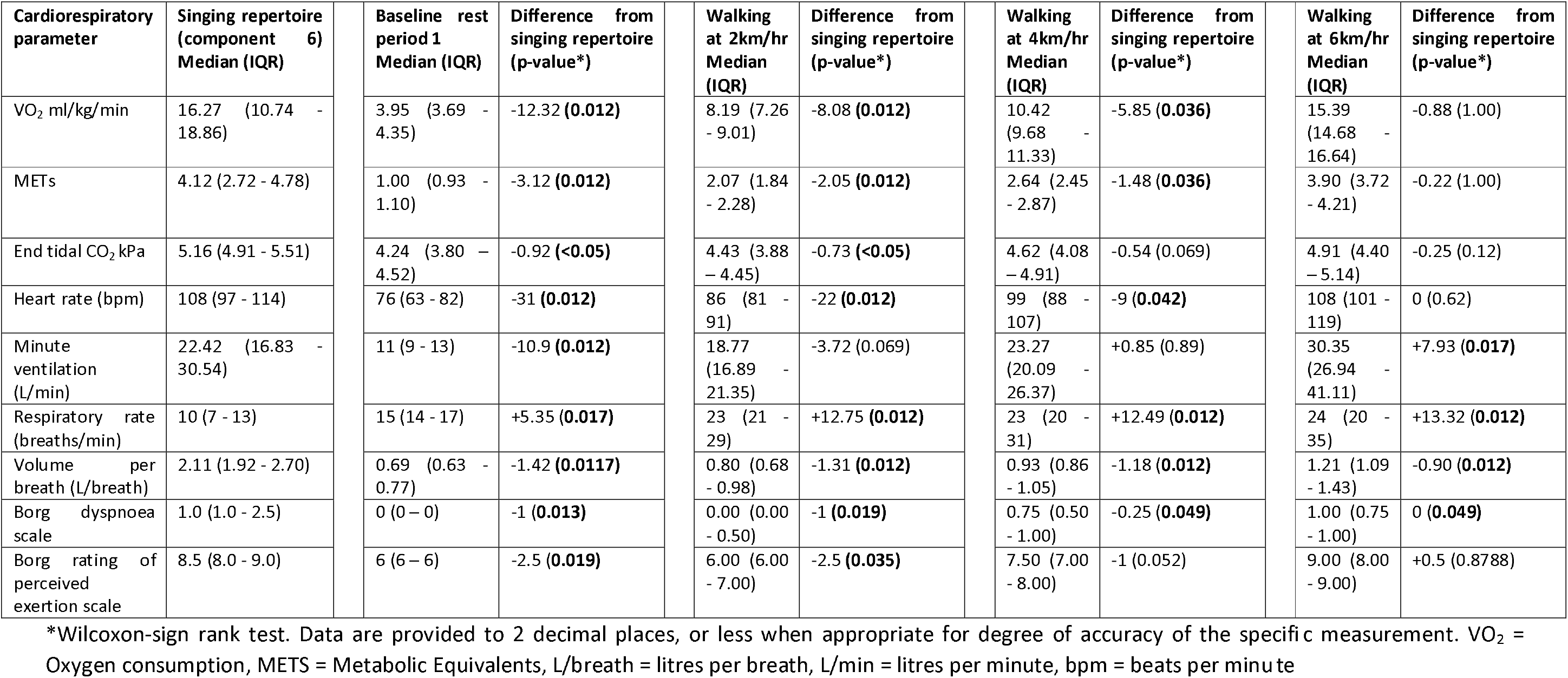
Comparison of singing with rest and walking at three different speeds.

| Cardiorespiratory parameter | Singing repertoire (component 6)<br>Median (IQR) | Baseline rest period 1<br>Median (IQR) | Difference from singing repertoire (p-value*) | Walking at 2km/hr<br>Median (IQR) | Difference from singing repertoire (p-value*) | Walking at 4km/hr<br>Median (IQR) | Difference from singing repertoire (p-value*) | Walking at 6km/hr<br>Median (IQR) | Difference from singing repertoire (p-value*) |
| --- | --- | --- | --- | --- | --- | --- | --- | --- | --- |
| VO<sub>2</sub> ml/kg/min | 16.27 (10.74 - 18.86) | 3.95 (3.69 - 4.35) | -12.32 (**0.012**) | 8.19 (7.26 - 9.01) | -8.08 (**0.012**) | 10.42 (9.68 - 11.33) | -5.85 (**0.036**) | 15.39 (14.68 - 16.64) | -0.88 (1.00) |
| METs | 4.12 (2.72 - 4.78) | 1.00 (0.93 - 1.10) | -3.12 (**0.012**) | 2.07 (1.84 - 2.28) | -2.05 (**0.012**) | 2.64 (2.45 - 2.87) | -1.48 (**0.036**) | 3.90 (3.72 - 4.21) | -0.22 (1.00) |
| End tidal CO<sub>2</sub> kPa | 5.16 (4.91 - 5.51) | 4.24 (3.80 - 4.52) | -0.92 (**<0.05**) | 4.43 (3.88 - 4.45) | -0.73 (**<0.05**) | 4.62 (4.08 - 4.91) | -0.54 (0.069) | 4.91 (4.40 - 5.14) | -0.25 (0.12) |
| Heart rate (bpm) | 108 (97 - 114) | 76 (63 - 82) | -31 (**0.012**) | 86 (81 - 91) | -22 (**0.012**) | 99 (88 - 107) | -9 (**0.042**) | 108 (101 - 119) | 0 (0.62) |
| Minute ventilation (L/min) | 22.42 (16.83 - 30.54) | 11 (9 - 13) | -10.9 (**0.012**) | 18.77 (16.89 - 21.35) | -3.72 (0.069) | 23.27 (20.09 - 26.37) | +0.85 (0.89) | 30.35 (26.94 - 41.11) | +7.93 (**0.017**) |
| Respiratory rate (breaths/min) | 10 (7 - 13) | 15 (14 - 17) | +5.35 (**0.017**) | 23 (21 - 29) | +12.75 (**0.012**) | 23 (20 - 31) | +12.49 (**0.012**) | 24 (20 - 35) | +13.32 (**0.012**) |
| Volume per breath (L/breath) | 2.11 (1.92 - 2.70) | 0.69 (0.63 - 0.77) | -1.42 (**0.0117**) | 0.80 (0.68 - 0.98) | -1.31 (**0.012**) | 0.93 (0.86 - 1.05) | -1.18 (**0.012**) | 1.21 (1.09 - 1.43) | -0.90 (**0.012**) |
| Borg dyspnoea scale | 1.0 (1.0 - 2.5) | 0 (0 - 0) | -1 (**0.013**) | 0.00 (0.00 - 0.50) | -1 (**0.019**) | 0.75 (0.50 - 1.00) | -0.25 (**0.049**) | 1.00 (0.75 - 1.00) | 0 (**0.049**) |
| Borg rating of perceived exertion scale | 8.5 (8.0 - 9.0) | 6 (6 - 6) | -2.5 (**0.019**) | 6.00 (6.00 - 7.00) | -2.5 (**0.035**) | 7.50 (7.00 - 8.00) | -1 (0.052) | 9.00 (8.00 - 9.00) | +0.5 (0.8788) |

\*Wilcoxon-sign rank test. Data are provided to 2 decimal places, or less when appropriate for degree of accuracy of the specific measurement. VO<sub>2</sub> = Oxygen consumption, METs = Metabolic Equivalents, L/breath = litres per breath, L/min = litres per minute, bpm = beats per minute

## Discussion

### Main finding

We found that the singing produced changes in physiological parameters including oxygen consumption, end tidal CO_2_, METs, heart rate, and minute ventilation, comparable to those seen when walking at a moderate to brisk pace, consistent with the changes in these parameters seen during moderate intensity physical activity.

Research regarding the oxygen cost of singing by non-professionals is limited. Sliiden et al (2016) present data from 20 musical theatre performers which suggest similar physiological responses to the current study when singing compared with rest, including heart rate, oxygen consumption, minute ventilation, and breath volume(30). Another study of nine musical theatre performers compared cardiorespiratory parameters while singing and dancing together, with dancing alone. The study found significantly lower breathing frequency and higher lactate when singing and dancing together, compared with dancing alone, but other parameters including oxygen consumption and heart rate did not differ significantly (31). However, singing alone (without dancing) was not compared to rest which limits comparisons. Regarding ventilatory volumes, our findings support previous research that suggest increases during singing and speech compared with spontaneous breathing (7, 32-36). However, much of the previous research concerns speech alone, and where singing has been investigated, the studies have largely focused on professional singers, or employed limited protocols that do not fully represent the range of activities engaged in during a community singing group. As such, application of previous research findings to the most common contexts in which people sing is challenging. To our knowledge this is the first study to systematically assess physiological parameters in amateurs, including pulmonary ventilation volumes, during the various singing activities commonly found in amateur community singing groups. As such, our findings build on those of other studies by demonstrating comparable physiological responses related to singing in non-professionals, and by comparing singing to a standardised form of physical activity in the form of treadmill walking.

Of note, the relative increases above baseline in ventilatory parameters may be of importance when considering aerosol transmission of infectious agents, including SARS-CoV-2, from both the perspective of people with the infection, and people who could be infected, in a setting. This is important particularly given that community singing groups have been identified as high risk for transmission(37, 38). Larger ventilatory volumes are relevant to dispersion of aerosols from infected individuals but may also impact the ‘dose’ of aerosols inhaled by those at risk of infection. As such, approaches such as remote singing groups delivered via video conferencing applications may mitigate associated risks, with pilot work suggesting potential health and wellbeing benefits from such approaches are still possible(18).

An important consideration when interpreting our findings, is that the extent to which people are moving is also likely to be a major factor in determining the physiological demands of the activity. Though completely static singing is unrealistic, we should consider that different types of singing encourage different levels of body movement, gesture, and dance like movements, in addition to voice production. Comparing the seated components of the protocol to standing singing (component 6), gives some indication of the contribution of posture and body movements to physiological demands.

A further point for consideration is the extent to which changes in the physiological parameters assessed result from physical exertion, or a degree of relative hyperventilation required for vocalisation. For example, one might expect to see larger ventilatory volumes, and possibly heart rate, because of the air flow velocity and volumes requirements for vocalisation. However, the pattern of end tidal CO_2_ during singing, compared with walking, suggests that hyperventilation alone does not account for the changes in the other parameters seen during the singing component. Furthermore, while minute ventilation approximately doubles from baseline, VO_2_ approximately quadruples, suggestive of an important contribution from higher cardiac output, respiratory muscle extraction, and skeletal muscles involved in movement, however the relative contribution of these factors has not been investigated here.

### Methodological considerations

This study has multiple strengths. To our knowledge, this is the first study to compare the physiological demands of singing to walking, using measures of ventilation, oxygen consumption, end tidal CO_2_, and perceived effort and dyspnoea simultaneously. The focus on amateur singers makes the findings highly relevant for the vast majority of people who sing.

Certain limitations should be mentioned. Firstly, the use of healthy, relatively young participants may limit the extent to which our findings can be extrapolated to older people, or those with significant medical conditions, such as those with chronic respiratory disease (CRD). However, individuals with CRD are likely to find activities such as singing, more rather than less physiologically demanding, as a proportion of their VO_2_ max(39). Therefore, one might reasonably suspect that the potential for physical benefits related to training effects would also be increased, though in what way, and to what extent, remains unclear. Secondly, the sample size is small; although it was sufficient to meet the aims of the study by comparing the parameters during protocol components, replication of our findings in larger samples is encouraged. Thirdly, although we considered real world applicability when developing the components of the protocol, the total protocol duration was approximately 25 minutes, while most community singing sessions are longer. As such, further studies during real world community singing group sessions would be of interest. Lastly, though this study has demonstrated that singing induces physiological responses that are similar in magnitude to moderate intensity physical activity, this study has not assessed training effects of singing. As such we cannot draw clear conclusions from this study alone regarding impacts on physical fitness.

To build on these findings, future research could include maximal exercise tests for comparison; explore the training effects following a programme of singing; directly compare professional and amateur singers; specifically assess the impact of musical genre, volume, and physical movements; and compare healthy controls with people with certain chronic diseases, in whom singing is being delivered in a therapeutic context.

## Conclusion

This study demonstrated that singing when standing induced physiological responses similar in magnitude to moderate intensity physical activity. The study also identified increases in minute ventilation, and breath volumes during singing and during singing related activities, that may be important when considering risk of transmission of respiratory infections including SARS-CoV-2. These findings suggest that health and wellbeing benefits attributed to singing participation, may in part, result from physical mechanisms. Further research including different types of singing, and singers, and training effects would be valuable.

## Supporting information

STROBE checklist

## Data Availability

Data may be made available on reasonable request.

## Funding statement

KP was supported by the Imperial College Clinician Investigator Scholarship. DF was supported by the Wellcome Trust [205407/Z/16/Z]. The funders had no say in the design and conduct of the study; collection, management, analysis, and interpretation of the data; preparation, review, or approval of the manuscript; and decision to submit the manuscript for publication.

## Competing Interests

No conflicts of interest declared.

## Authors Contributions

KP and AL had the original idea for the study, and collected the data with assistance of CM. KP analysed the data and wrote the first draft of the manuscript. All authors contributed to the study design, writing, reviewing and editing the manuscript, and approved the final manuscript for submission.

## Acknowledgements

We would like to give our thanks to the study participants for their time and effort, and Tim Grove who reviewed physiological aspects of the manuscript.

KP would like to acknowledge the National Institute for Health Research (NIHR) Biomedical Research Centre based at Imperial College Healthcare NHS Trust and Imperial College London for their support. The views expressed are those of the authors and not necessarily those of the NHS, the NIHR or the Department of Health.

## Data Availability Statement

Data may be made available on reasonable request.

## Supplement

### 1. Component learning phase

Consenting participants took part in SLH activities under the instruction of Dr Adam Lewis (AL) who teaches on the Singing for Lung Health Singing Leader training programmes and demonstrated component exercises during the set up and calibration of other equipment. These simple exercises are regularly undertaken by people with significant medical problems, so we anticipated that all participants would have been able to complete these activities. However, participants were informed they were free to stop at any point. Participants had to demonstrate, repeat and confirm understanding of the example exercises of each component demonstrated, and AL had to be satisfied they could be perform them effectively, keeping to the rhythm of AL’s performance for each component. Participants were instructed that they would be led throughout the protocol by AL and able to mirror the activities shown. Participants were given the opportunity to practice and ask questions about each component Learning all the singing activities took approximately 15 minutes. All participants had previously used the treadmill equipment during other studies and so no learning phase was required for this.

The timing of each following component and instruction period was recorded by KP.

Component 1: Rest period 1, baseline assessment: Participants sat for five minutes at rest. Two minutes of resting condition was then recorded prior to taking part in six of the core components of a SLH session for periods of two minutes per component.

Component 2: Physical warm up: gentle, gestural, dance-based movements, standing, performed to music (“Sugar Sugar”, by The Honeys): Participants commenced a physical warm up accompanied to music: The song was played via Adam Lewis’s mobile phone. Participants followed AL through the following in sitting: Alternate toe taps, Alternate heel digs, hamstring curls under chair, alternate punching then increasing range of flexion (pretending to climb a ladder), forward leaning and sweeping the floor with both hands moving all the way to full shoulder flexion with widely spread hands (all movements should be comfortably within the participants individual range of movement). The participants were then instructed to stand and repeat in standing followed by neck rotation and side-flexion exercises. Participants returned to a sitting position at the end of the two minute period.

Component 3: Singing based rhythm exercise. Participants were instructed to follow AL through a song called ‘Alive, Awake, Alert, Enthusiastic’ This phrase was repeated with each word having an associated body action: Both AL and participants touched their heads with both hands for ‘Alive’, touched their shoulders for ‘Awake’, touched their knees for ‘Alert’ and clapped, then flex their shoulders to 90 degrees and had elbows fully extended and wrists supinated for ‘enthusiastic’. These actions were repeated in song. Individuals then dropped the ‘Alert’ action and sung word. This was then repeated by singing ‘Alert’ but dropping ‘Enthusiastic’. The Rhythm exercise was be performed in a sitting posture. Participants were instructed to get each repetition of ‘Alive, awake, alert, enthusiastic’ sung in a single breath.

Component 4: Pitch exercise: seated singing, focusing on challenging the accuracy and range of pitch achieved. Participants were instructed to follow AL singing a song with the lyrics: ‘Elevator won’t you take me 1,2,3,4,5. Elevator won’t you take me 5,4,3,2,1’ This phrase was repeated with a higher pitch being reached with each consecutive number and the pitch coming back down in scale with each descending number. Participants mirrored AL in standing, with feet slightly wider than shoulder width, with slight flexion in the knees. With each increasing number that was sung, the participant raised their left hand parallel to their xiphisternum (‘1’) in approximately 10cm increments until level ‘5’ was approximately at the level of their nose. The phrase was repeated with number ‘3’ being absent and then repeated with both number ‘3’ and ‘5’ being absent. All numbers were then sung again and repeated. Participants were encouraged to get the whole sung phrase out in 1 breath.

Component 5: Fricatives – a vocal exercise focusing on consonant vocalisations. Participants were asked to mirror AL through a set of voiced fricatives in a standing posture in step stance. ‘Ssshhhh’ ‘jjjjjj’ and ‘vvvv’ were repeated with participants putting one hand on their abdomen just above the pelvic bone, and the other hand below the xiphisternum. Then alternating hand position by both hands moving to be in a fist position and pushed into their sides in between the lower ribs and hips. These hand positions were designed to provide tactile feedback from the abdominal muscle use with the exhaled breath. Participants were then asked to pulse the voiced fricatives in a ‘1, 2’ rhythm. Finally participants were asked to repeat the voiced fricatives whilst in alternate step stance.

Component 6: Repertoire: Singing standing up. Participants sang ‘1 bottle of beer’ acapella. The lyrics are as follows:

> One bottle of beer, two bottle of beer, three bottle of beer,
>
> four bottle of beer, five bottle of beer, six bottle of beer,
>
> seven bottle of beer, 8 POP! (1 breath)
>
> Fish and chips and vinegar, vinegar, vinegar,
>
> Fish and chips and vinegar, pepper, pepper, pepper POT! (1 breath)
>
> Oh you can’t put your muck in our dustbin,
>
> our dustbin, our dustbin,
>
> You can’t put your muck in our dustbin,
>
> our dustbin’s FULL! (1 breath)

Individuals were encouraged to repeat each segment in one exhaled breath. Individuals were asked to count using their hands and fingers for actions and sway laterally with each number with feet slightly wider than shoulder width and slightly flexed knees. This swaying motion is exaggerated with ‘fish and chips…’ pretending to cradle the fish and chips like a baby as if they were wrapped in newspaper. Then during the ‘muck in our dustbin’ participants will be asked to stamp alternate feet forward and point with alternate hands repeatedly.

Component 7: Rest component 2: This was included to i) compare with the baseline measurements, to assess if the protocol included sufficient time for full recovery between components; and ii) to enable to participants to return to baseline before the walking components. This was an active rest period as it was accompanied by some relaxation prompts given by AL. Participants were instructed to sit down and follow a guided visualisation relaxation with imagery of being on a beach relaxing on a deck chair with the warm sun on the face and the perfect breeze flowing and the smooth rhythmical sound of waves in the background. Instructions regarding optimising individual’s body posture were given and any points of muscular tension that was noted by AL was addressed with guided muscle relaxation instructions such as ‘releasing the jaw’, and ‘open your palms to the sky’.

Component 8: treadmill walking at 2km/hr, no incline

Component 9: treadmill walking at 4km/hr, no incline

Component 10: treadmill walking at 6km/hr, no incline

## Additional Graphs

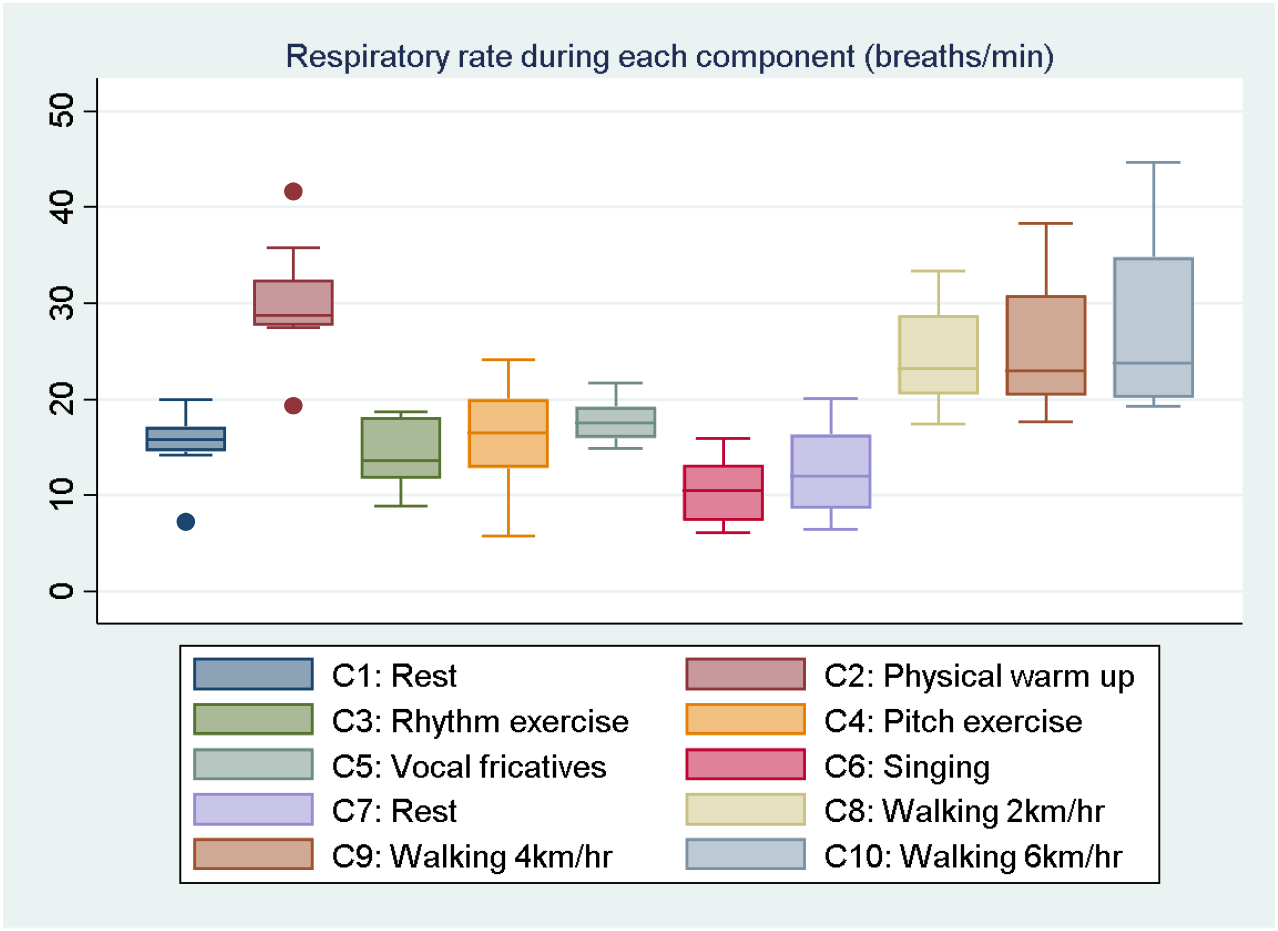

## Notes

### Competing Interest Statement

The authors have declared no competing interest.

### Clinical Trial

NCT04121351

### Author Declarations

Ethical approval was granted by the Imperial College Research Ethics Committee (IREC) (19IC5429). All participants provided informed written consent.

